# Saturation-scale functional evidence supports clinical variant interpretation in Lynch Syndrome

**DOI:** 10.1101/2022.08.08.22278549

**Authors:** Anthony Scott, Felicia Hernandez, Adam Chamberlin, Cathy Smith, Rachid Karam, Jacob O. Kitzman

## Abstract

Lynch Syndrome (LS) is a cancer predisposition syndrome affecting more than 1 in every 300 individuals worldwide. Clinical genetic testing for LS can be life-saving but is complicated by the heavy burden of variants of uncertain significance (VUS), especially missense changes. To address this challenge, we leveraged a multiplex analysis of variant effect (MAVE) map covering >94% of the 17,746 possible missense variants in the key LS gene *MSH2*. Here, to establish the clinical validity of these functional data, and to demonstrate their utility in large-scale variant reclassification, we overlaid them on clinical databases comprising >15,000 individuals with an MMR gene variant uncovered during clinical genetic testing. Our functional measurements agreed with the clinical interpretation for every one of 47 control variants with available classifications, satisfying accepted thresholds for ‘strong’ evidence for or against pathogenicity. We then used these scores to attempt reclassification for 682 unique missense VUS, among which 34 (5.0%) scored as deleterious in our function map, in line with previously published rates among other cancer predisposition genes. Consistent with their pathogenicity, functionally abnormal missense variants were associated with significantly elevated risk for LS-related cancers. Combining functional data and other lines of evidence, ten variants were reclassified as pathogenic/likely pathogenic, and 497 could be moved to benign/likely benign. Finally, we applied these functional scores to paired tumor-normal genetic tests, and identified a subset of patients with biallelic somatic loss of function, reflecting a sporadic Lynch-like Syndrome with distinct implications for treatment and relatives’ risk. This study demonstrates how high-throughput functional assays can empower scalable VUS resolution and prospectively generate strong evidence for variant classification.

## Introduction

Identification of a pathogenic variant in a familial cancer risk gene can inform treatment and prevention strategies for patients and their blood relatives. As a prominent example of a common and broadly screened cancer risk syndrome, Lynch Syndrome (LS) affects nearly 1 in 300 individuals worldwide [1,2], and is primarily associated with colorectal and endometrial cancers. Heterozygous carriers’ risk for these cancers approaches ∼80% and ∼60%, respectively [3,4], with onset decades earlier on average compared to sporadic cases [5].

LS is caused by inherited defects in any of four key DNA mismatch repair (MMR) factors: *MSH2, MLH1, MSH6*, and *PMS2*. These genes are included on most cancer gene panel tests, and it is standard of care to screen them for pathogenic germline variants when their loss is observed in tumors by histology or by tests for microsatellite instability, the mutational consequence of MMR loss. Increased carrier screening in LS holds great potential for reducing risk: it is estimated that the large majority of individuals who carry an LS variant go undetected at present [6], in part because many of them lack the clear family history needed to meet current genetic testing criteria [7,8].

Despite decades of clinical screening and functional studies, upwards of one-third of the variants identified during clinical genetic evaluation cannot be classified, and remain as variants of uncertain significance, or VUS [9,10]. The eventual reclassification rate for VUS in LS and other hereditary cancer genes is modest, reaching only ∼25% [11,12]. The difficulty of resolving these VUS stands as one of the most persistent barriers to the utility of broader genetic testing for LS genes [13].

As a group, missense variants are a particularly challenging to interpret as most are individually rare (i.e., population minor allele frequencies ≤10^−4^), and they can have functional defects resulting from diverse molecular mechanisms [14]. Over 94% (n=8,614) of the LS gene missense variants in the NCBI ClinVar database [15] remain as VUS or have conflicting interpretations, reflecting both the volume of VUS’ discovery and the challenge of their classification. Even when tumor molecular testing is available, it may not resolve these variants’ effects – for example, missense variants can retain protein staining by immunohistochemical testing despite causing mismatch repair deficiency, a source of false negatives that could potentially limit access to immunotherapy [16].

Functionally testing individual missense VUSs in experimental model systems is time- and labor-intensive but can provide key evidence to guide their classification. New approaches (collectively termed multiplex assays of variant effect, or MAVEs) have enabled the systematic testing of many variants at a time [17,18]. Promisingly, benchmarking comparisons have shown that MAVEs can accurately recapitulate standing classifications from sources such as ClinVar [19,20]. However, despite the recent proliferation of MAVE studies, practical examples of their use in clinical variant interpretation have been scarce. One recent study [21] evaluated MAVE scores’ predictive accuracy for variants found during genetic testing in three cancer-associated genes (*BRCA1, TP53*, and *PTEN*). Under recently proposed guidelines [22,23], the authors were able to reclassify just over half (176/324) of the VUSs that had functional information across the three genes. In another recent study, the same *BRCA1* MAVE scores [24] were intersected with variants discovered during exome sequencing of an unselected healthcare cohort, and an association between MAVE-predicted loss of function variants and breast and ovarian cancer diagnoses was observed [25].

Here, we set out to use MAVE-based function maps to facilitate variant classification in Lynch Syndrome. We combined a recent MAVE [26] covering 16,749 missense variants in the key Lynch Syndrome gene *MSH2* with a clinical dataset containing 15,520 patients with an LS gene variant. We validated the *MSH2* MAVE data across 47 previously classified missense variants, and found that it meets the established threshold for ‘strong’ functional evidence [27]. Critically, during validation, we excluded any variants for which the standing classification relied upon functional data, thus avoiding the risk of circularity inherent in validating one functional assay using another. We then applied these MAVE scores to 682 standing MSH2 missense VUS, formally reclassifying 10 to pathogenic/likely pathogenic, and showing that another 497 could be moved to benign/likely benign upon reassessment. Leveraging the detailed, individual-level clinical information in this cohort, we demonstrate that missense *MSH2* variants with abnormal MAVE function scores are associated with elevated colorectal and endometrial cancer risk. Finally, going beyond germline variant interpretation, we demonstrate that MAVE scores can uncover loss-of-function somatic ‘second hits’ as well as biallelic mutations with the availability of tumor DNA tests.

## Methods

### Patient population

Clinical information and genetic variants were obtained for patients found to carry at least one variant in any of the four major Lynch Syndrome genes (*MSH2, MLH1, MSH6, PMS2*) during multi-gene panel testing for cancer predisposition at Ambry Genetics before December 14, 2020. We obtained data for 13,916 LS gene variant carriers who underwent germline-only testing, and another 1,604 patients who underwent paired tumor-germline testing at Ambry Genetics before August 31, 2020. Data collection and sharing procedures were reviewed by the Western IRB and University of Michigan Medical School IRB (study HUM00220511) and declared exempt from human subjects regulations.

### Functional annotation of *MSH2* missense variants

Each *MSH2* missense variant was annotated with two function scores: the loss of function (LoF) scores from a recent deep mutational scan [26] which measures impact upon MSH2 protein function, and SpliceAI deltaMax scores [28], a computational estimate of the probability of splicing disruption. Variants with an LoF score ≥ 0.4, or a SpliceAI deltaMax score ≥0.5 were considered deleterious; those with LoF scores between 0 and 0.4, or deltaMax between 0.2 and 0.5 were considered intermediate, while those with LoF scores < 0 and deltaMax < 0.2 were considered functionally neutral.

### Variant classification

Patient variant classification was performed at Ambry Genetics using a points-based implementation [29] of ACMG/AMP variant classification guidelines [23,30], assigning each variant into one of five tiers: pathogenic (P), likely pathogenic (LP), uncertain significance (VUS), likely benign (LB) or benign (B). To validate the *MSH2* missense function scores, we used previously classified *MSH2* missense variants from the paired (tumor-normal) dataset. To avoid redundant application of evidence, we used only those variants which had sufficient evidence to be classified as benign/likely benign and pathogenic/likely pathogenic without use of any prior functional data. Function scores’ strength of evidence for or against pathogenicity was quantified using the Oddspath score [22,31]. Additionally, structural evidence was assessed using a standard structural modelling protocol and energies of destabilization compared to nearby informative variants and identification of impacted motifs. (Martin et al. 2017, Sherrill et al. 2018).

### Cancer association

For analysis of cancer prevalence among LS variant carriers, patients with cancer were categorized by variant classification(s) and gene(s) affected. For analysis of *MSH2* VUS carriers, we excluded individuals who also carried a pathogenic or likely pathogenic (P/LP) variant in a non-Lynch Syndrome gene, while individuals with both an *MSH2* VUS and a P/LP variant in *MLH1, MSH6*, or *PMS2* were considered as carriers for those respective genes and were excluded from *MSH2* VUS association tests. Logistic regression models were fit using the python stats models package (version 0.12.2), using cancer diagnosis as the response variable and, as features, each individuals’ carrier status for the following categories of variants, each encoded as zero or one: (1) *MSH2* missense with deleterious function score, (2) MSH2 missense with neutral function score, (3) *MSH2* other P/LP, (4) *MLH1* any P/LP, (5) *MSH6* any P/LP, and (6) *PMS2* any P/LP. Models were fit separately for colorectal cancer and uterine/endometrial cancer (in the latter case, including only females).

## Results

### Clinical validation of MSH2 function map

To establish the clinical validity of multiplexed analyses of variant effect (MAVEs) for interpreting and reclassifying variants in Lynch Syndrome, we intersected loss-of-function (LoF) scores from a deep mutational scan of *MSH2* [26] with a clinical database of 1,604 individuals with MMR gene variants detected by paired tumor and germline testing. To gauge the strength of evidence provided by our functional data, we curated a list of *MSH2* missense variants previously classified as pathogenic/likely pathogenic (n=22) or benign/likely benign (n=26) as known controls. Crucially, to avoid the circularity of validating one functional assay using classifications that relied upon other mechanistically similar assays, we included only variants for which clinical interpretation could be reached without the use of any prior functional evidence (e.g., only using population frequency, family history, or tumor characteristics).

Our functional measurements agreed with the clinical interpretation of all 47 of the 47 variants that scored as functionally abnormal or normal (**Figure 1**, and **Supplementary Table 1**), with one pathogenic variant scoring in the intermediate range. This resulted in a strength of evidence, as quantified by the OddsPath score [31], of 24.9 for abnormal LoF scores, and 0.043 for neutral scores. Following recommendations for application of the functional evidence criterion using the ACMG/AMP variant interpretation framework [27], *MSH2* LoF scores ≥ 0.4 can therefore be used as “strong evidence” in support of variant pathogenicity (PS3 evidence code), while LoF scores < 0 can be conversely be used a “strong evidence” against pathogenicity (BS3 evidence code).

**Figure 1.**
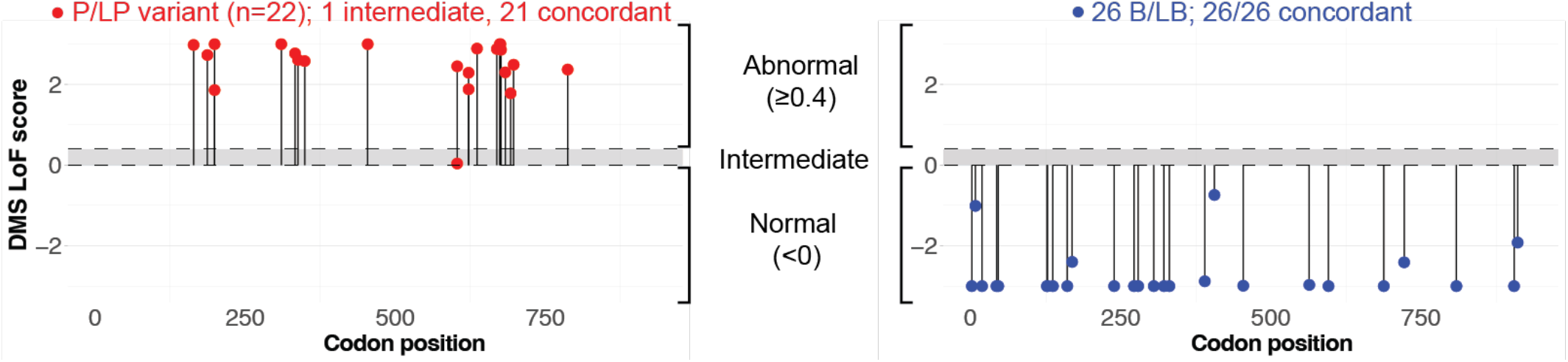
Validation of *MSH2* function scores across 48 previously classified *MSH2* missense variants. LoF scores for known pathogenic/likely pathogenic (red, at left) and known benign/likely benign variants (blue, at right) are plotted against codon position. Gray shaded interval denotes intermediate score range.

We next applied the *MSH2* missense function scores to a larger panel of individuals (n=13,916) for whom germline-only testing had identified at least one germline Lynch Syndrome gene variant. Among this cohort, 1,937 individuals carried a scorable *MSH2* missense variant. We focused first on the 32 distinct missense variants, carried by 108 individuals, which had been previously classified by the clinical laboratory as pathogenic or likely pathogenic. Of those, 31 had an abnormal function score. The lone exception was the missense variant c.2020G>A (p.Gly674Ser), which was classified as likely pathogenic and has been shown to be partially deficient in ATP binding *in vitro* [32]; this variant may be a false negative in the functional assay. Of the 31 correctly identified pathogenic variants, 27/31 had an abnormal LoF score from deep mutational scanning, while 4/31 were predicted to be splice disruptive by SpliceAI, indicating that at least among known variants in *MSH2*, disruption at the level of protein function contributes a greater share of the pathogenic burden than splicing defects. In all, the *MSH2* missense function scores achieved a recall of 96.9% in a validation dataset independent from the patient cohort used to derive the OddsPath score.

For the overwhelming majority of *MSH2* missense carriers in this cohort, the variants carried were VUS (1,829 individuals, 87.4%). Of the 682 unique such missense VUS, 5.0% scored as functionally abnormal: 24 by DMS LoF score and another 10 by SpliceAI (with another 17 in the intermediate range by either measure; **Figure 2**). Thus, loss of function was modestly depleted among these extant variants, relative to the 5,130 missense SNVs not observed in this study, among which 6.8% were abnormal (298 and 53 each by DMS and SpliceAI, respectively; P=0.048, two-sided binomial test). The depletion of functionally deleterious variants among standing missense VUSs likely reflects the ongoing removal of those with sufficient lines of evidence to be classified as pathogenic.

**Figure 2.**
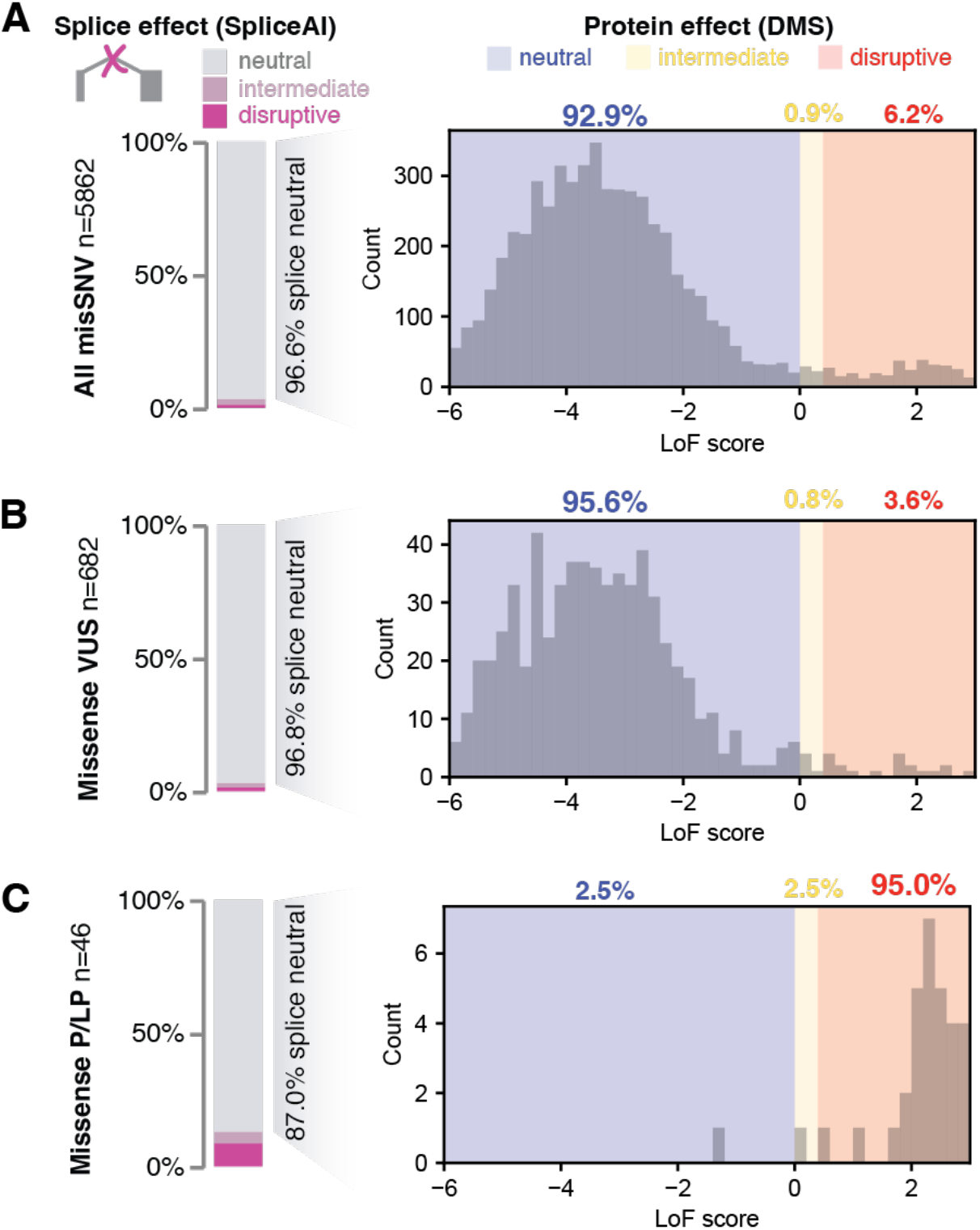
Function scores and variant reclassification for *MSH2* missense variants, for **(a)** all single nucleotide missense variants, **(b)** missense VUSs, and **(c)** missense variants previously classified as pathogenic or likely pathogenic, including those used for validation. For each group of variants, splicing status was scored by SpliceAI (bar charts at left), and for splice-neutral variants (SpliceAI score<0.2), a histogram of LoF scores from deep mutational scanning are displayed to the right

### VUS reclassification

We next pursued clinical variant reclassification for *MSH2* missense variants with abnormal DMS LoF scores, adding functional evidence codes in support of their pathogenicity. Of the 24 such variants, 14 variants had scores exceeding (i.e., more abnormal) the lowest score in the P/LP validation set (LoF score ≥1.7), and we added PS3 (strong evidence) codes for each of these. After adding this evidence, ten of these VUSs met criteria to be reclassified as pathogenic or likely pathogenic (**Table 1**). Consistent with their causal, pathogenic role, nine of these ten exhibited MMR deficiency as shown by IHC and/or MSI testing. To be conservative, we did not pursue formal reclassification for variants with LoF scores between 0.4-1.7; while these were in the abnormal range, they scored below all the training variants used to establish the OddsPath score, and these could in principle be given a weaker evidence code (PS3_moderate) in the future. In sum, there were 14 remaining missense VUSs in the functionally abnormal range which require additional evidence for formal reclassification under the ACMG/AMP framework, including observation in additional cases, or co-occurrence with a somatic loss-of-function variant in the same gene. In the other direction, there were 635 patient missense VUSs (carried by 1,772 individuals) which were functionally normal by both deep mutational scanning and SpliceAI prediction. We set out to determine what percentage could potentially be reclassified as benign/likely benign by adding a BS3 functional evidence code and found that 497 of these variants (78.3%) could be reclassified to B/LB with the addition of that evidence. Thus, with the addition of functional evidence, approximately three-quarters of all standing *MSH2* missense VUS in this large patient cohort could be newly classified (**Figure 3**).

**Table 1.** Summary of reclassification for missense *MSH2* germline VUS with abnormal LoF scores (n=24), with evidence codes applied for each variant. *PS3_moderate codes are applicable for variants with scores between 0.4 and 1.7, but their reclassification was not pursued here.

| Variant | Reclassification Outcome | LOF Score (Jia et al. 2021) | MAPP MMR(C hao et al. 2008) | gnom AD frequency | LOF score evidence | Pop. database evidence | Clinical (MSI/IHC) | Computational evidence | Additional Evidence |
| --- | --- | --- | --- | --- | --- | --- | --- | --- | --- |
| V695M | Reclassified to LP | 4.511 | 20.15 | - | PS3 | PM2_P | PP4 |  |  |
| L310R | Reclassified to LP | 3.774 | 33.02 | - | PS3 | PM2_P | PP4_S | PP3 | PP1 |
| F523S | Reclassified to LP | 3.547 | 10.09 | - | PS3 | PM2_P | PP4 |  | PM1 |
| V148E | Insufficient evidence | 3.239 | 4.13 | - | PS3 | PM2_P | PP4 |  |  |
| N671Y | Reclassified to LP | 3.114 | 20.2 | - | PS3 | PM2_P | PS4_M | PP3 | PM1_P |
| D603V | Reclassified to LP | 2.999 | 22.25 | - | PS3 | PM2_P | PP4 | PP3 |  |
| I356K | Reclassified to LP | 2.518 | 14.66 | - | PS3 | PM2_P | PP4 | PP3 |  |
| T677R | Reclassified to LP | 2.469 | 37.76 | - | PS3 | PM2_P | PP4_M | PP3 | PM1 |
| G759V | Insufficient evidence | 2.215 | 29.36 | - | PS3 | PM2_P |  | PP3 |  |
| G761R | Reclassified to LP | 2.193 | 40.89 | - | PS3 | PM2_P |  |  | PM1_P |
| D758Y | Insufficient evidence | 1.955 | 15.52 | - | PS3 | PM2_P |  |  |  |
| A700E | Reclassified to LP | 1.917 | 37.91 | - | PS3 | PM2_P | PP4 |  |  |
| G761V | Insufficient evidence | 1.785 | 9.19 | - | PS3 | PM2_P |  |  |  |
| L414P | Reclassified to LP | 1.766 | 12.66 | - | PS3 | PM2_P | PP4 |  | PM1 |
| V923E | Conflicting/insufficient evidence | 1.669 | n/a | - | PS3_moderate* | PM2_P | BS2_P |  |  |
| T552P | Insufficient evidence | 1.61 | 10.84 | - | PS3_moderate* | PM2_P | PS4_M |  |  |
| R534P | Insufficient evidence | 1.383 | 30.22 | - | PS3_moderate* | PM2_P |  |  |  |
| T568I | Conflicting/insufficient evidence | 0.932 | 4.04 | - | PS3_moderate* | PM2_P |  | BP4 |  |
| R621P | Insufficient evidence | 0.745 | 30.86 | - | PS3_moderate* | PM2_P | PP4 |  |  |
| D758H | Insufficient evidence | 0.701 | 18.79 | - | PS3_moderate* | PM2_P |  |  |  |
| S129C | Conflicting/insufficient evidence | 0.574 | 1.84 | 2.35E-05 | PS3_moderate* |  | PP4 |  | BS2_P |
| A309P | Insufficient evidence | 0.528 | 7.68 | - | PS3_moderate* | PM2_P |  | PP3 |  |
| R711L | Conflicting/insufficient evidence | 0.491 | 19.03 | - | PS3_moderate* | PM2_P | BS2_P | PP3 |  |
| R524L | Insufficient evidence | 0.449 | 17.81 | - | PS3_moderate* | PM2_P |  |  |  |

**Figure 3.**
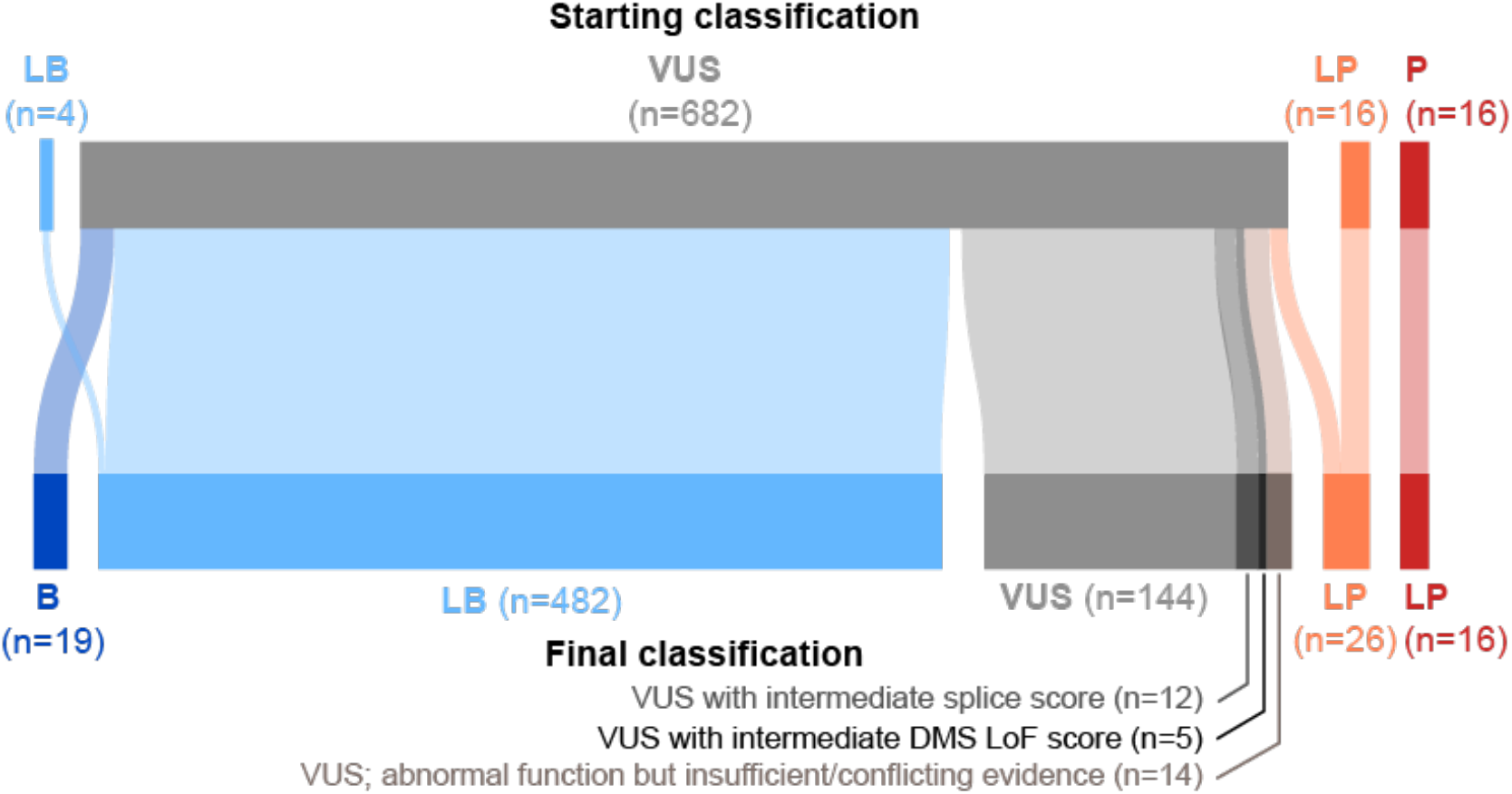
Reclassification outcomes for 718 *MSH2* missense variants. Flow diagram showing starting and final variant classifications; in total, 76% of the missense VUSs have sufficient evidence to enable potential reclassification to benign (B), likely benign (LB), likely pathogenic (LP) or pathogenic (P). A subset of remaining VUSs had intermediate function scores (n=17), or had abnormal function scores but lacked sufficient lines of evidence (or had conflicting evidence) and so remain as VUS (n=12).

### Cancer prevalence among LOF VUS missense carriers

We next sought to compare the risk conferred by LOF *MSH2* missense variants to that of established P/LP variants in *MSH2* and other LS genes. In this patient cohort, LS-related cancer diagnoses were enriched relative to the general population, but far from completely prevalent: 13.6% of patients in this cohort had a CRC diagnosis, with higher rates in males (38.3%, n=2,229) compared to females (8.9%, n=11,687), possibly reflecting broader inclusion criteria for genetic screening in women (e.g., for breast cancer). Uterine and endometrial cancers (UEC) were similarly prevalent to CRC, affecting 9.5% of females. Other cancers not primarily associated with Lynch Syndrome were also prevalent in this cohort, affecting 49.6% of females and 40.7% in males. Gene by gene differences in penetrance closely mirrored those seen previously [2,33]: P/LP variants in *MLH1* and *MSH2* were the most strongly associated with CRC (odds ratio=14.4 and 8.10, respectively), with lesser effects from *MSH6* and *PMS2* P/LP variants. As previously noted [5], uterine and endometrial cancers differed from colorectal cancer, with *MSH6* (OR=13.2) and *MSH2* (OR=11.9) emerging as the top risk factors, followed by *MLH1* and *PMS2*. Separating *MSH2* missense variants by their functional status, those with abnormal function scores (by DMS or SpliceAI) were significantly associated with both CRC (OR=2.53, 95%CI:[1.04, 6.15], P=0.04) and EC (OR=5.56, 95%CI:[2.24,13.8], P=2.2×10^−4^), though with smaller effects than truncating P/LP variants’ (**Figure 4**). By contrast, *MSH2* missense variants with neutral function scores did not contribute significant risk for CRC or EC (P≥0.67 for each), nor were they associated with other cancers (**Supplementary Figure 1**). Thus, loss-of-function missense variants in MMR genes contribute measurable risk for LS-associated cancers, but may exhibit lower risk than their truncating counterparts, underscoring the challenge of their accurate classification.

**Figure 4.**
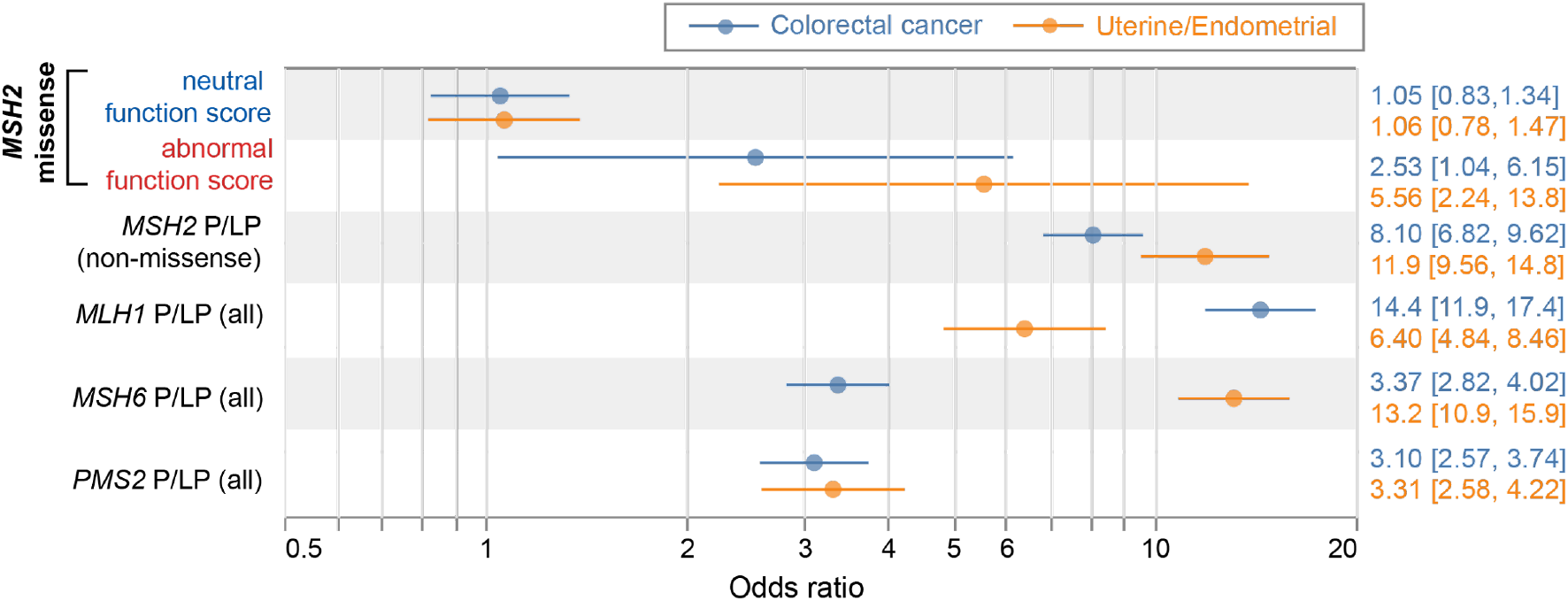
Cancer associations by variant type. Association between colorectal cancer (blue) or uterine/endometrial cancer (female) and missense variants in *MSH2* (missense, separated by DMS+SpliceAI function score), or P/LP variants in other Lynch Syndrome genes; odds ratios shown from logistic regression.

### Joint annotation of germline and somatic variants

We next sought to apply these functional measures to jointly interpret germline and somatic mutations in *MSH2*. Most cases of Lynch Syndrome follow a ‘two hit’ model, with one inherited loss-of-function variant followed by a second somatic mutation disrupting the remaining copy. Therefore, it is expected that in a cohort including individuals with LS, pathogenic somatic ‘second hits’ in *MSH2* would be more common among those individuals who inherited a ‘first hit’ loss-of-function variant in the same gene. We tested this within the paired tumor-normal cohort (n=1,282 individuals), among the 25 individuals for whom the sole germline finding was a missense *MSH2* variant (**Figure 5** and **Supplementary Table 2**). DMS scores indicated 13 of these 25 germline variants are functionally deleterious, constituting pathogenic inherited ‘first hits’. Among these 13 carriers, 12 (92.4%) had a P/LP somatic ‘second hit’ in *MSH2*, or a structural variant in the upstream gene *EPCAM* (which causes epigenetic silencing of *MSH2* [34,35]. By contrast, among the other 12 individuals who inherited a single *MSH2* missense variant scored as neutral by DMS, only two (16.7%) had a P/LP somatic mutation in *MSH2*, a significantly lower prevalence (P=0.00021, Fisher’s Exact test).

**Figure 5.**
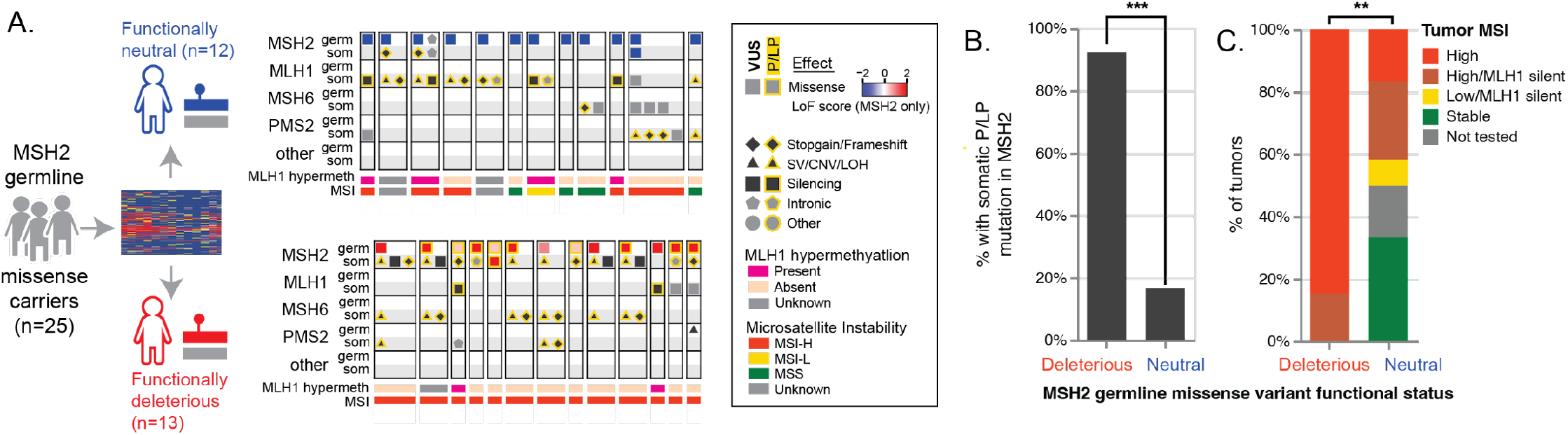
Joint analysis of germline and tumor mutations. **(A)** Patterns of germline and somatic mutations among LS genes, among germline carriers of *MSH2* missense variants, separated into those scored as functionally neutral (top) or deleterious (bottom) by deep mutational scanning LoF score **(B)** Fraction of individuals with a somatic P/LP mutation in *MSH2*, by *MSH2* germline missense functional status. (**C)** Tumor microsatellite status, by *MSH2* germline missense variant functional status. ***, P<0.001; **, P<0.01

Tumor microsatellite instability (MSI), a hallmark of MMR deficiency, was universal among patients with functionally abnormal *MSH2* missense alleles. After excluding individuals with *MLH1* promoter hypermethylation (an independent somatic epigenetic mechanism sufficient to cause MMR deficiency), tumors from all 11 of 11 individuals who carried a germline *MSH2* missense variant deemed LoF by DMS were MSI-high, whereas only 2/6 individuals with a germline functionally neutral missense *MSH2* variant had MSI-high or MSI-low tumors (P=0.006), and both of those two cases could be explained by biallelic somatic mutations in other LS genes (PMS2:c.1687C>T:p.R563* + LOH in one, MLH1:c.676C>T:p.R226* + LOH in the other). The ability of the *MSH2* function map to identify germline variants associated with pathogenic somatic second hits and MSI demonstrates how patterns of tumor mutation can support germline variant classification [36].

Somatic mutations also contribute to the variant interpretation burden, particularly for heavily mutated MMR-deficient tumors. Indeed, among the 437 individuals for which paired testing revealed at least one somatic *MSH2* mutation, most (382, 87.4%) had more than one somatic mutation in a tested gene, and nearly half (182, 47.6%) had multiple somatic mutations in *MSH2* alone. We focused on the 84 individuals who carried at least one missense somatic *MSH2* variant, with 46 carrying at least one somatic missense *MSH2* variant we predict to be functionally disruptive, and 38 for whom these somatic missense variants were exclusively functionally neutral and without any other somatic P/LP *MSH2* mutations (**Figure 6 and Supplementary Table 3**). Notably, among the latter (carriers of exclusively functionally neutral somatic *MSH2* variants), additional somatic mutations in other LS genes (i.e., *MSH6, MLH1, PMS2*) were found in all 38 tumors (100%). By contrast, when at least one functionally disruptive *MSH2* missense somatic mutation was found, somatic mutations in other LS genes were significantly less common (30 of 46 tumors; Fisher’s exact P=2.70×10^−5^). Similarly, *MLH1* promoter hypermethylation was present in nearly a third of the tumors for which the only somatic *MSH2* mutations were functionally neutral missense (9 of the 28 tumors in which *MLH1* was assayed, 32.1%) but nearly absent among tumors with at least one somatic missense *MSH2* mutation deemed LoF by DMS/SpliceAI (1 of 38, 2.6%; Fisher’s exact P=0.0013). MAVE measurements can thus identify functionally disruptive somatic mutations driving tumor MMR deficiency even in the absence of an inherited loss of function variant.

**Figure 6.**
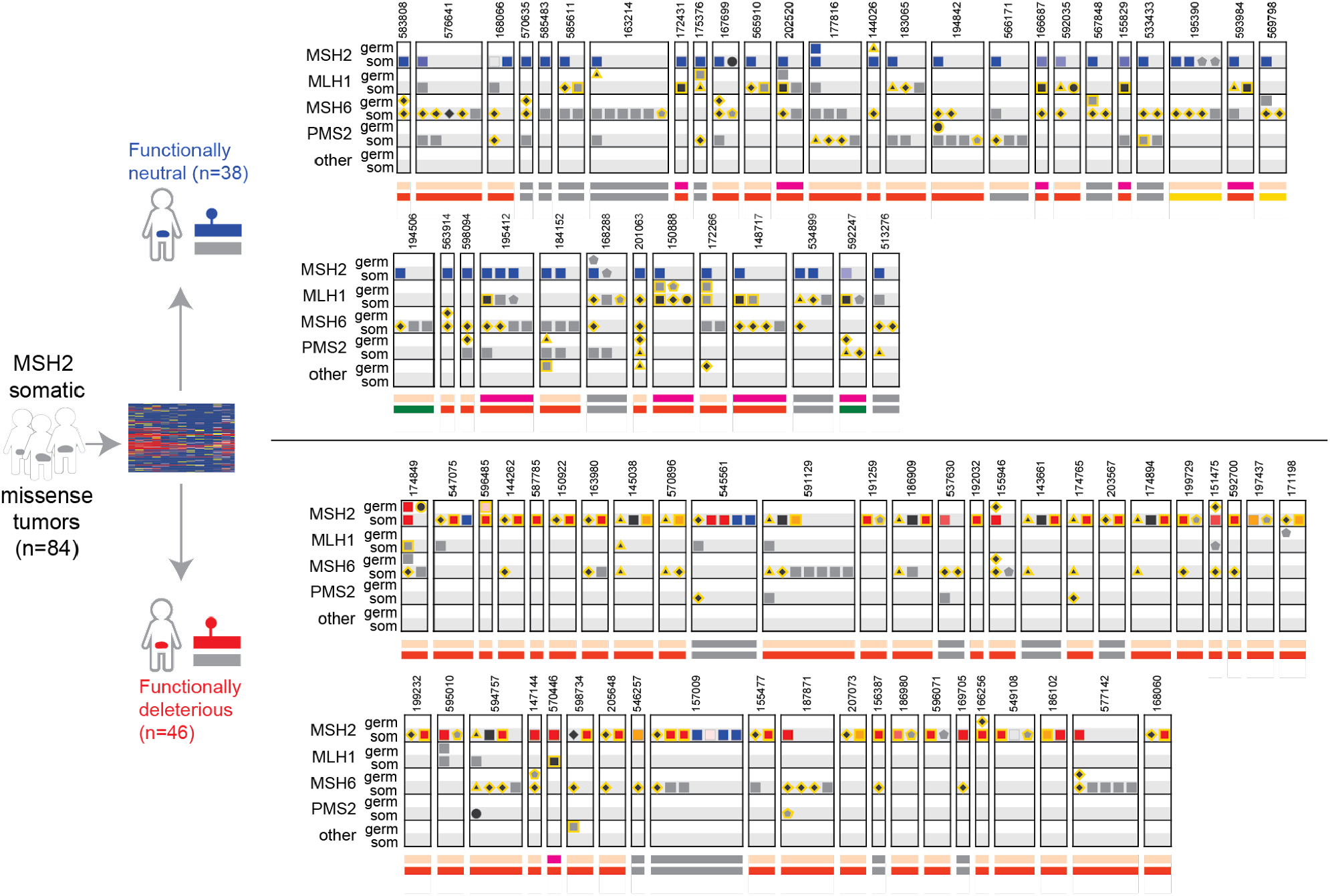
Mutational patterns in patients with somatic *MSH2* missense variants. Tumor and germline mutations in LS genes are shown in patients who carry functionally neutral somatic *MSH2* missense variants (upper track, n=38), or those with a functionally abnormal somatic *MSH2* variant (lower track, n=46). Mutations and tumor characteristics are denoted as in Figure 5A.

## Discussion

As MAVE function maps are put into practice for clinical variant interpretation, an important prerequisite is to assess their predictive value for disease risk and clinical phenotypes. Here we establish the validity of MAVE-based functional evidence for missense variant classification in the Lynch Syndrome gene *MSH2*. We supplemented protein-level MAVE effect measurements with deep learning-based splicing effect predictions [28], to newly reclassify over 74% of the 682 missense variants of uncertain significance (VUS) encountered in *MSH2*, in a cohort of tens of thousands of individuals with genetic testing results.

In particular, the reclassification of 10 variants to pathogenic/likely pathogenic newly enabled the return of definitive genetic diagnoses. Reclassification of these variants as pathogenic has critical clinical implications for these patients and family members who also inherited them, such as more frequent colonoscopies, risk-reducing surgeries to avoid gynecologic cancers, initiation of esophagogastroduodenoscopy for upper GI cancer surveillance and additional cancer screening recommendations not necessarily performed for the general population. Going forward, these MAVE-based function scores are now integrated into the variant interpretation process at clinical genetic testing laboratories, and will assist with classification of newly observed rare *MSH2* missense variants.

Our study leverages several unique features of this large cohort. Firstly, to validate the *MSH2* MAVE, we selected a set of control variants for which the classification stands without including prior functional evidence, that is, based upon orthogonal features such as recurrence, tumor characteristics, and co-segregation with early-onset cancer. This avoids the risk of validating a MAVE in part by prior functional evidence from existing assays, which despite being lower-throughput, may be mechanistically similar and highly correlated with the MAVE.

For many genes, culling the training set to remove such variants may not be practical -- obtaining a sufficient number of control variants is emerging as a key rate limiting step for many MAVEs; at least 11 control variants are needed to reach a ‘moderate’ strength of evidence [27]. This challenge is highlighted by a recent effort to reclassify variants in *PTEN* using MAVE data [21], which was hampered by the limited number (n=2) of known benign variants. In many cases, filtering variants for this or other criteria may not even be possible: public-facing databases such as ClinVar are often the primary source for these controls, and to protect privacy they do not provide individual-level clinical or demographic data.

We used per-individual clinical information to explore the association between loss-of-function, as indicated by MAVE scores, and cancer prevalence. We observed that *MSH2* missense variants with abnormal function identified by MAVE were associated with significantly elevated cancer risk for LS-associated colorectal and uterine/endometrial cancers. Notably, these associations were weaker than those observed for P/LP variants that were not missense (i.e., truncating frameshift or stop-gain variants). Thus, functionally abnormal *MSH2* missense variants as a group may be less penetrant than their truncating counterparts, while still being measurably pathogenic within the population of individuals selected for germline cancer testing.

In contrast, another recent study of MMR gene variant carriers found no difference in the incidence of LS-related cancers between carriers of *MSH2* missense P/LP and truncating variants [37]. A key difference, however, was that the missense variants included in that study were restricted to those with standing P/LP classifications, which may reflect a particularly severe subset. Thus, the addition of MAVE-based functional data may have captured missense variants with intermediate functional defects which confer a moderate level of risk. An important future direction will be to replicate this analysis in an unselected population, as has recently been done for *BRCA1* [25], and to model polygenic risk as a potential modifier [38].

To date, applications of MAVE data have largely focused on germline variants. Here, we demonstrated how MAVEs can also support joint analyses of germline and somatic mutations, leveraging a clinical database of 1,282 individuals with paired tumor-normal tests. As expected under Knudson’s two-hit hypothesis [39], among individuals who inherited an *MSH2* missense variant, pathogenic somatic ‘second hits’ in *MSH2* were significantly more common when the MAVE data indicated the germline variant was functionally disruptive as compared to normal. In addition, we identified 29 individuals whose cancers had double *MSH2* somatic mutations with at least one of the mutations identified as functionally deleterious by MAVE. Excluding an inherited predisposition as the cause for these individuals’ MMRd tumors has the potential to prevent unnecessary screenings for their blood relatives.

A limitation of this study is that the MAVE function scores used here were derived from a cDNA-based deep mutational scan and so do not capture splice disruptive effects, which although in the minority relative to protein-disruptive variants, may still account for a substantial number of cases for LS [40,41]. These effects can be obtained experimentally with other MAVE approaches such as saturation genome editing [24] or saturation prime editing [42], or directly measured by massively parallel splicing assays [43–45]. For the purposes of this study, we used predictions from SpliceAI [28], a deep learning-based splicing effect predictor which has been shown to be highly accurate [46].

As gene panel and exome sequencing are increasingly utilized in the clinical setting for a variety of indications, there is an opportunity to leverage the massive scale of MAVE experiments to prospectively generate functional evidence for as yet unseen variants. With proper clinical validation, it appears promising that MAVE data may soon play a primary role in identifying patients who may not have otherwise come to clinical attention, but could benefit from additional monitoring based upon their genetic risk.

## Data Availability

All data produced in the present study are available upon reasonable request to the authors

## Acknowledgements

We thank Matthew Varga and Min-Sun Park of Ambry Genetics for support of the structural investigation of these variants, and members of the Kitzman lab for helpful comments.

## Funding

This work was supported by the National Institute of General Medical Sciences (R01GM129123 to J.O.K.).

## Author contributions

F.H. and R.K. abstracted clinical information. A.S., F.H., A.C., C.S., and J.O.K. analyzed the data. A.S., F.H., R.K., and J.O.K. wrote the manuscript.

## Competing interests

F.H., A.C., and R.K. are employees of Ambry Genetics. J.O.K. serves as a scientific advisor to MyOme, Inc. The authors declare that there are no further competing interests.

## Notes

### Author Declarations

The University of Michigan Medical School IRB reviewed this study (#HUM00220511) and waived ethical approval.

