## Supplementary figures and images for "Saturation-scale functional evidence supports clinical variant interpretation in Lynch Syndrome"

### Supplementary Figure 1

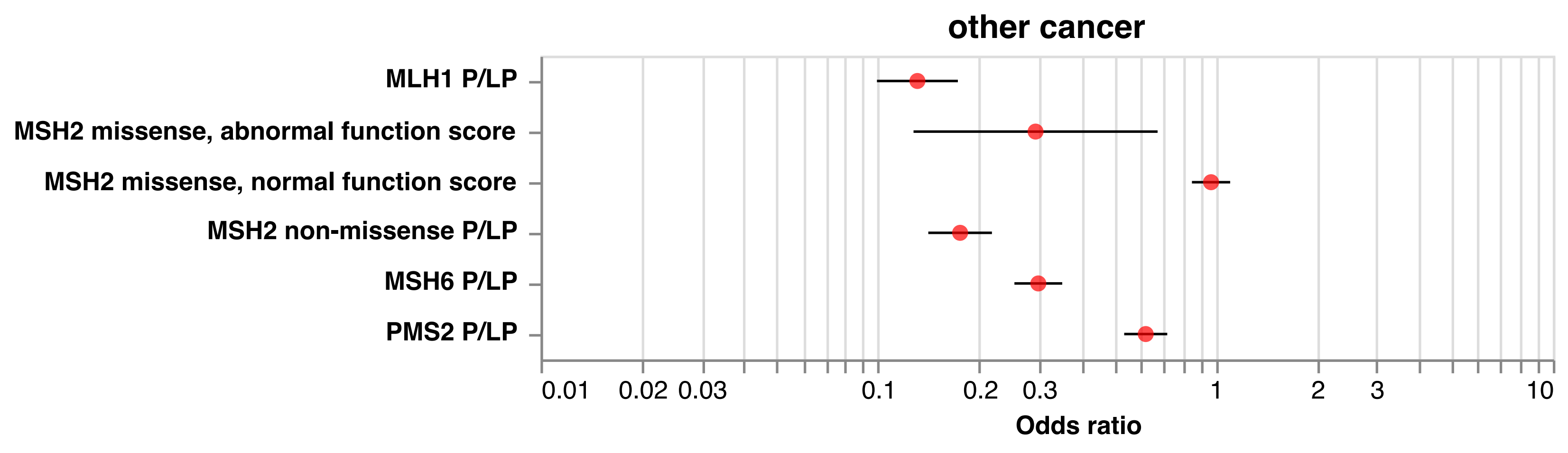
